# Adaptive immunity to human coronaviruses is widespread but low in magnitude

**DOI:** 10.1101/2021.01.24.21250074

**Authors:** Hyon-Xhi Tan, Wen Shi Lee, Kathleen M Wragg, Christina Nelson, Robyn Esterbauer, Hannah G Kelly, Thakshila Amarasena, Robert Jones, Graham Starkey, Bao Zhong Wang, Osamu Yoshino, Thomas Tiang, M Lindsay Grayson, Helen Opdam, Rohit D’Costa, Angela Vago, The Austin Liver Transplant Perfusionist Group, Laura K Mackay, Claire L Gordon, Adam K Wheatley, Stephen J Kent, Jennifer A Juno

## Abstract

Endemic human coronaviruses (hCoV) circulate worldwide but cause minimal mortality. Although seroconversion to hCoV is near ubiquitous during childhood, little is known about hCoV-specific T cell memory in adults. We quantified CD4 T cell and antibody responses to hCoV spike antigens in 42 SARS-CoV-2 uninfected individuals. T cell responses were widespread within conventional memory and cTFH compartments but did not correlate with IgG titres. SARS-CoV-2 cross-reactive T cells were observed in 48% of participants and correlated with HKU1 memory. hCoV-specific T cells exhibited a CCR6^+^ central memory phenotype in the blood, but were enriched for frequency and CXCR3 expression in human lung draining lymph nodes. Overall, hCoV-specific humoral and cellular memory are independently maintained, with a shared phenotype existing among coronavirus-specific CD4 T cells. This understanding of endemic coronavirus immunity provides insight into the homeostatic maintenance of immune responses that are likely to be critical components of protection against SARS-CoV-2.

## Introduction

In contrast to the high pathogenicity of MERS-CoV, SARS-CoV and SARS-CoV-2 coronaviruses, endemic human coronaviruses (hCoV) circulate worldwide but typically cause common colds with only limited morbidity and mortality^1^. Endemic hCoV encompass two alpha-coronaviruses (αCoV), NL63 and 229E, and two beta-coronaviruses (βCoV), HKU1 and OC43^1^. Sero-epidemiological studies suggest that infection and seroconversion to hCoV occurs during early childhood (typically by 4 years of age)^2-4^, although there are discrepant reports on the prevalence of each virus within distinct geographical cohorts^2,5^. Despite the early development of immunity against multiple hCoV, most adults remain susceptible to periodic reinfection^6-8^, with increased susceptibility among immunocompromised individuals^9-11^. This suggests the magnitude and/or quality of hCoV-targeted immunity in adults is insufficient for sterilizing protection but instead may limit the burden of disease to asymptomatic or mild infection^8^. Defining the extent of serological and/or cellular immunity required to protect individuals from reinfection or severe disease remains a key question in the SARS-CoV-2 pandemic. As neutralizing responses wane after CoV infection, it is likely that a combination of serum antibody and B cell / T cell memory provide longer-term protection from the recurrence of disease^12,13^. The study of hCoV-specific T and B cell memory can therefore provide a key preview into the development of durable, protective SARS-CoV-2 immunity.

Characterisation of population-level immunity to hCoV can also inform our understanding of cross-reactive immune responses between high pathogenicity and endemic CoV. Studies of SARS-CoV-2-specific immunity in uninfected individuals clearly demonstrate pre-existing cross-reactive antibody^14-16^, B cell^16^ and T cell responses^17-22^. Nevertheless, it is currently unclear what contribution, if any, cross-reactive immunity plays in modulating the response to SARS-CoV-2 infection or vaccination^23^. Detailed analyses of cross-reactive T cells suggest the majority of such responses are dominated by CD4 T cells and directed toward non-RBD epitopes of the spike (S) protein^18,21,24^. To date, however, consensus regarding the origin of these cross-reactive responses is lacking, with evidence both for^18^ and against^25^ a major contribution from hCoV-specific memory T cells.

Deconvolution of cross-reactive SARS-CoV-2 responses and de novo SARS-CoV-2 immunity requires a more detailed understanding of hCoV-specific serological and cellular memory. Relatively little is known about population-level T or B cell memory to hCoV in adults, despite evidence suggesting an impact of recent hCoV infection on COVID-19 severity^26^. Several groups find widespread but modest CD4 T cell responses to hCoV proteins, with estimates for the prevalence of memory responses ranging from 70-100% of study participants^24,25,27^. Detection of hCoV-specific CD8 T cell responses has been less reported^27^, and the prevalence of cross-reactive SARS-CoV-2-specific responses in these cohorts varies substantially^24,25^. Furthermore, data comparing hCoV-specific T or B cell responses in the circulation with the presence or absence of such responses in the respiratory tract or secondary lymphoid organs (SLO) is lacking. Studies in animal models suggest that respiratory infections can generate long-lived T cell memory in lung draining lymph nodes (LDLN)^28^, raising the possibility of analogous responses following hCoV infection.

To address these knowledge gaps, we assessed the prevalence and phenotypic characteristics of hCoV spike-specific antibody, memory T cell and memory B cell responses in a cohort of SARS-CoV-2 uninfected adults. We find that the magnitude of hCoV immunity is independent of age and is characterized by robust antibody titres, widespread CD4 T cell memory within both Tmem and cTFH populations, and an enrichment of T cell memory in LDLN. In contrast, neutralizing antibody activity is relatively low and memory B cells are infrequently detected in either the circulation or LDLN. Overall, our data detail a consistent pattern of hCoV-specific immune memory in the circulation and SLO which likely co-ordinate to provide long-term protection from hCoV infection.

## Results

### hCoV-specific antibody and CD4 T cell memory is common among adults

We recruited a cohort of 42 SARS-CoV-2 uninfected adults (n=21 male, n=21 female), ranging in age from 18-67 years with no recent cold or COVID-19 symptoms (Figure 1A). Consistent with previous studies^15,16^, we detected baseline plasma antibody responses to one or more hCoV S antigens in all participants, with substantially lower reactivity toward SARS-CoV-2 S (herein CoV-2; Figure 1B). Plasma IgG endpoint titres for hCoV antigens ranged from 1:176 to 1:18268 (median 1:1485 for HKU1, IQR 1:886.8-2045; median 1:4475 for OC43, IQR 1:2082-6979; median 1:2066 for 229E, IQR 1:1185-3789; median 1:1716 for NL63, IQR 1:1193-2731).

**Figure 1.**
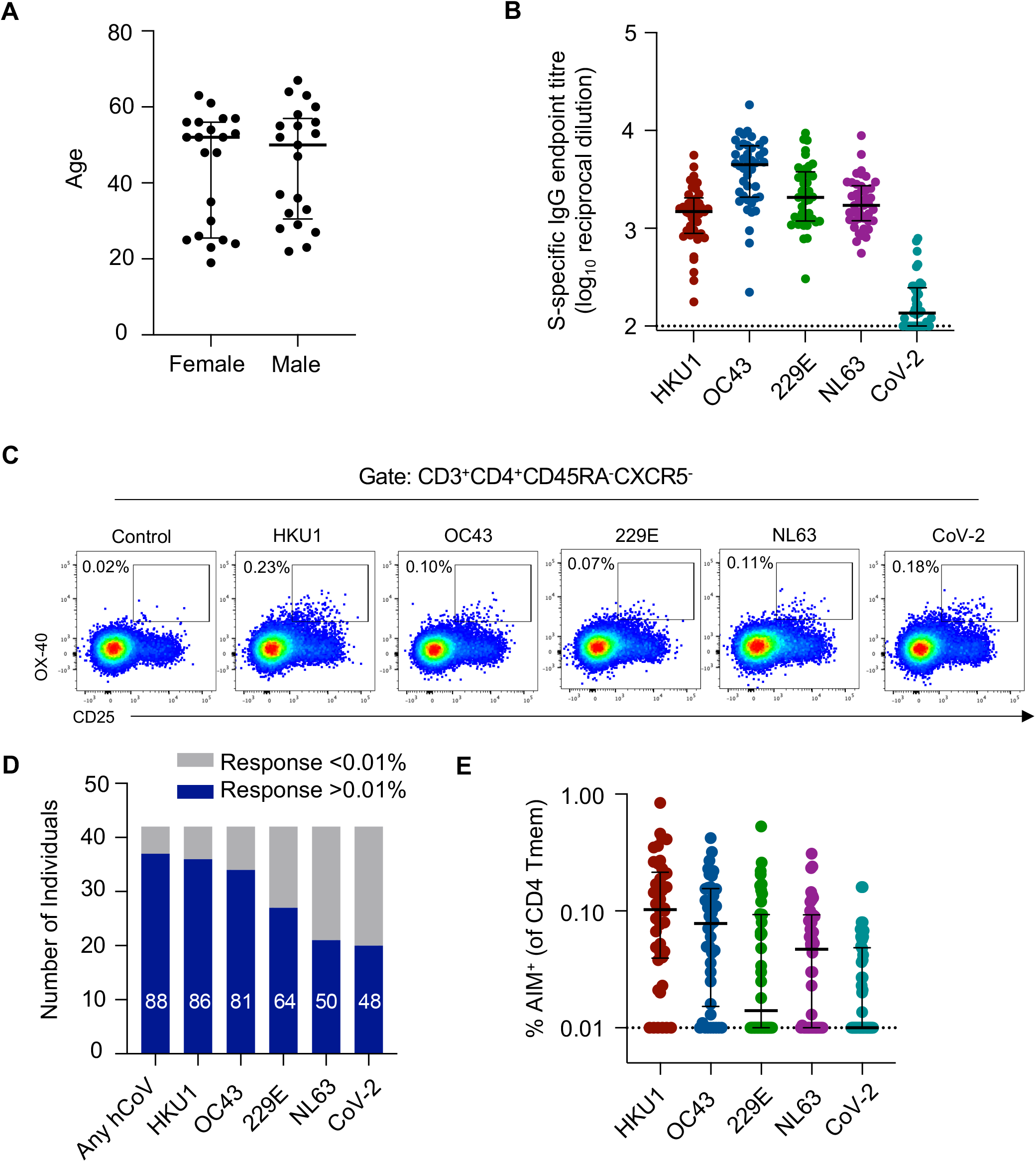
hCoV and CoV-2 CD4 Tmem responses among healthy subjects. (A) Ages of the CoV-2 uninfected adult cohort participants (n=21 female, n=21 male) (B) Plasma samples were screened by ELISA for reactivity against S proteins from hCoV or CoV-2 (n=42). Data are presented as the reciprocal endpoint titre. Dashed line indicates the limit of detection of the assay. (C) Representative plots of coronavirus S-specific CD4 Tmem (CD3^+^CD4^+^CXCR5^-^ CD45RA^-^) from a single individual measured by OX-40 and CD25 expression (control well stimulated with BSA). (D) Number of individuals with S-specific responses greater than 0.01% of total Tmem for each indicated antigen (n=42). Numbers in bars indicate the percentage of responders for each antigen. (E) Frequency of S-specific Tmem for each antigen (n=42). Lines indicate median. Values represent background subtracted responses; frequencies below 0.01% after background subtraction were assigned a value of 0.01%.

To determine the distribution of CD4 T cell memory responses, we stimulated PBMC with recombinant S antigens and quantified antigen-specific Tmem (CD3^+^CD4^+^CD45RA^+^CXCR5^-^) by measuring upregulation of the activation markers CD25 and OX-40 by flow cytometry (a well-established activation-induced marker (AIM) assay^29-31^) (Figure 1C; gating in Supplemental Figure 1). Across the cohort, 88% of individuals exhibited a memory response greater than 0.01% above background^32^ to any hCoV S antigen (Figure 1D). Interestingly, the prevalence of responses was highest to HKU1 S (86% of participants), and lowest to NL63, with only 50% of individuals exhibiting NL63 S-specific responses (Figure 1D). The magnitude of responses to hCoV S antigens ranged from undetectable to a maximum of 0.84% of the Tmem compartment (Figure 1E). Among individuals with above-background responses, median antigen-specific Tmem frequencies were highest to HKU1 (median 0.133%, IQR 0.056-0.248, n=36), followed by OC43 (median 0.106%, IQR 0.049-0.170, n=34), NL63 (median 0.093%, IQR 0.055-0.168, n=21), and 229E (median 0.080%, IQR 0.050-0.124, n=27). Similar to other cohorts^17,19^, we find 48% of participants (n=20) demonstrated cross-reactive response to CoV-2 S with a median frequency of 0.049% (IQR 0.027-0.160), despite no evidence of prior infection (Figure 1D/E). T cell responses were similar when measured using either CD25/OX-40 or CD137/OX-40^19^ AIM assays (Supplementary Figure 2A-C). Across the cohort, there was no relationship between the total frequency of hCoV S-specific Tmem and age, or any association with gender (Supplementary Figure 3A-B).

### hCoV-specific CD4 Tmem are predominately T_CM_ cells with a CCR6^+^ phenotype

Given divergent host receptor specificity and possible differences in tissue tropism among hCoV^1^, we assessed whether memory or chemokine receptor phenotypes differed among S-specific CD4 T cell populations (gating in Supplementary Figure 1B). Similar to the parental Tmem population, hCoV S-specific and CoV-2 cross-reactive CD4 T cells were predominately CD27^+^CCR7^+^, classically defined as central memory T cells (T_CM_; Figure 2A-B). In contrast to the bulk Tmem population, however, hCoV S-specific cells were substantially enriched for CCR6 expression (with or without co-expression of CXCR3; Figure 2C-D). When comparing intra-individual responses, hCoV S-specific Tmem phenotypes were generally similar across all S antigens (Figure 2E). Prior studies have also described a dominant CCR6 phenotype of CoV-2 S-specific Tmem among convalescent COVID-19 subjects^33^, and here we find that CoV-2 cross-reactive responses are similarly highly CCR6 biased (Figure 2D-E).

**Figure 2.**
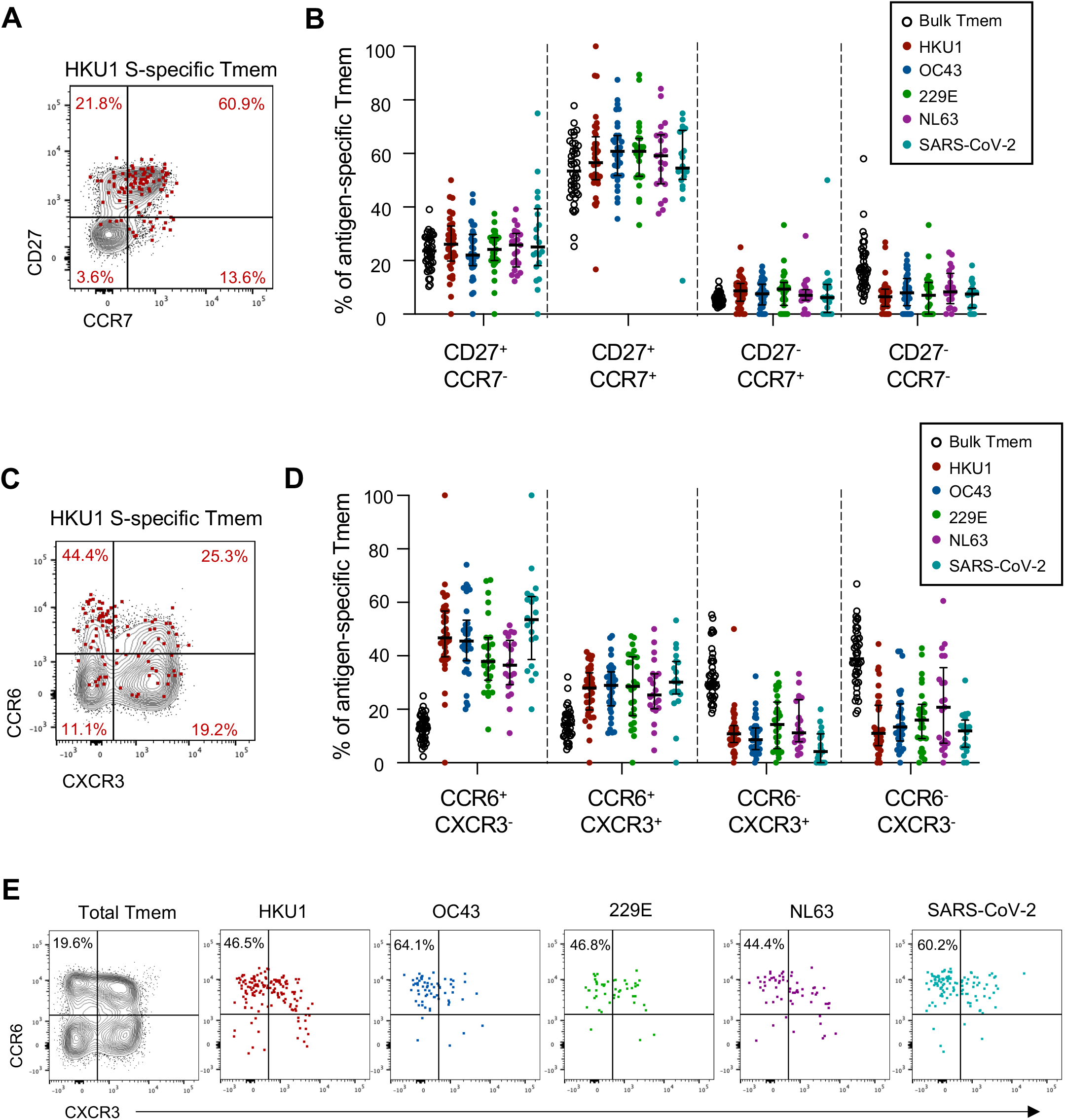
Memory and Th phenotype of hCoV and cross-reactive SARS-CoV-2 CD4 Tmem responses. (A) Representative staining of CD27 and CCR7 on the total Tmem population (black) or HKU1 S-specific Tmem (red) in a single donor. (B) Quantification of memory phenotype among bulk Tmem (n=42), HKU1 (n=36), OC43 (n=34), 229E (n=27), NL63 (n=21) or CoV-2 (n=20)-specific Tmem. (C) Representative staining of CCR6 and CXCR3 on the total Tmem population (black) or HKU1 S-specific Tmem (red) in a single donor. (D) Quantification of Th phenotype among bulk Tmem (n=42), HKU1 (n=36), OC43 (n=34), 229E (n=27), NL63 (n=21) or CoV-2 (n=20)-specific Tmem. (E) Comparison of hCoV and SARS-CoV-2-specific T cell phenotype in a single donor with responses to all antigens. In all graphs, individuals were excluded if they did not exhibit a response to a particular antigen.

### hCoV reactivity is detected among circulating T follicular helper cell (TFH) memory

Circulating TFH cells (cTFH; CXCR5^+^CD45RA^-^) comprise a clonally^34^ and functionally^29,35^ distinct memory CD4 T cell population identified by CXCR5 expression. Activated cTFH correlate with antibody responses to infection or vaccination, and are thought to be surrogates of germinal centre (GC) TFH activity^36,37^. Resting cTFH, in contrast, may represent a long-lived, homeostatic memory population from which recall responses can be elicited even years after antigen exposure^38-41^. Like conventional Tmem, hCoV-specific and cross-reactive cTFH responses were widely detected across the cohort (Figure 3A). The frequency of donors exhibiting cTFH responses above 0.01% to each antigen was similar to that observed for Tmem responses (90% for HKU1, 88% for OC43, 69% for 229E, 59% for NL63, 43% for CoV-2). Median frequencies among responding donors were highest to HKU1 (median 0.241%, IQR 0.147-0.531), followed by OC43 (median 0.213%, IQR 0.126-0.424), 229E (median 0.126%, IQR 0.061-0.340), NL63 (median 0.096%, IQR 0.050-0.210) and CoV-2 (median 0.085%, IQR 0.050-0.195) (Figure 3A).

**Figure 3.**
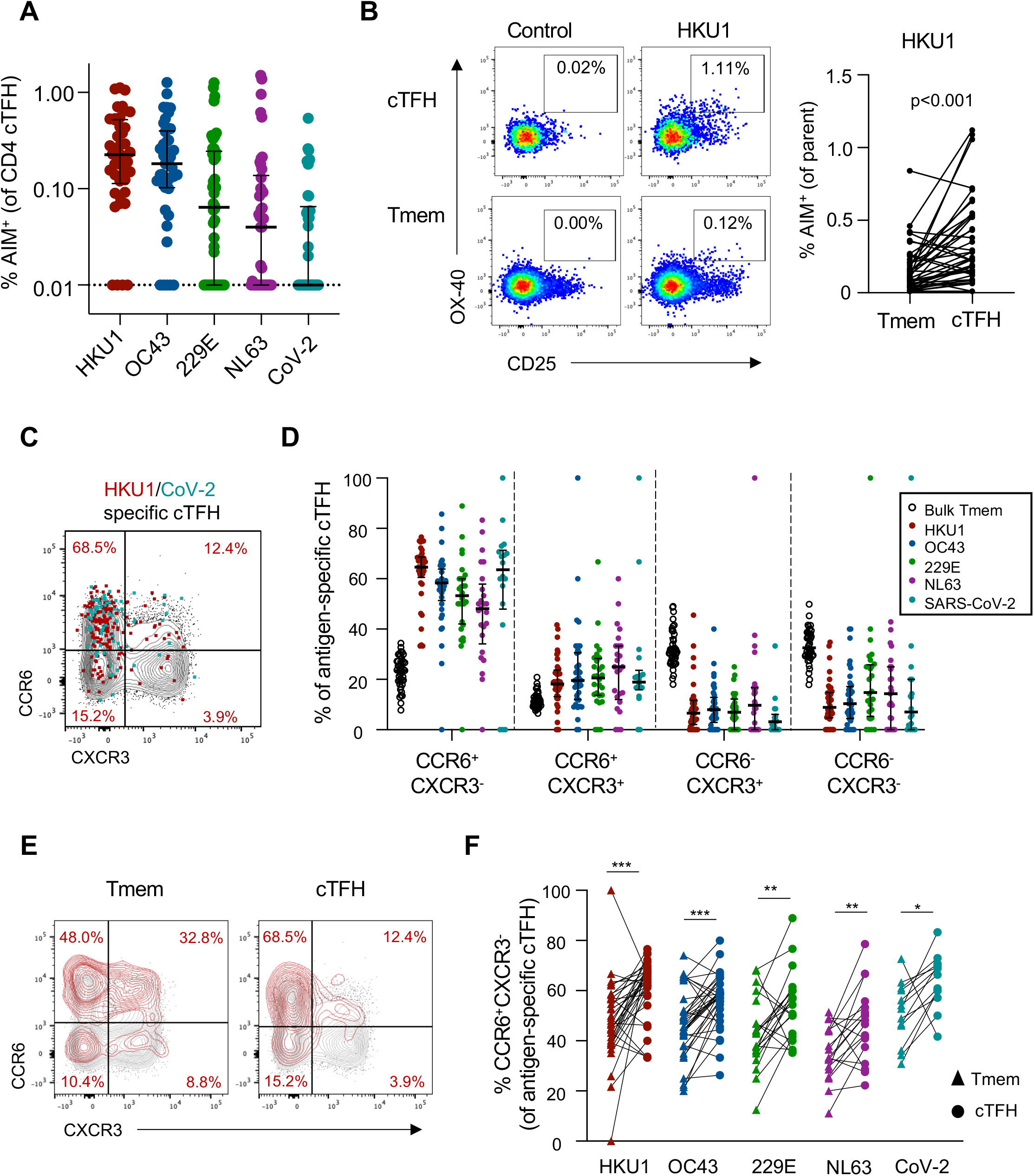
Memory and Th phenotype of hCoV and cross-reactive CoV-2 cTFH responses. (A) Frequency of S-specific cTFH for each antigen (n=42). Lines indicate median. Values represent background subtracted responses; frequencies below 0.01% after background subtraction were assigned a value of 0.01%. Data points are segregated and coded as individuals without (closed circles, n=24) or with (open circles, n=18) CoV-2 cross-reactive responses. Comparison of HKU1 S-specific T cell frequencies in Tmem or cTFH subsets. Plots indicate representative data from one donor. Graph shows compilation of responses from all donors (n=42). (C) Representative staining of CCR6 and CXCR3 on the total cTFH population (black), HKU1-specific (red) or CoV-2-specific cTFH (teal) in a single donor. (D) Quantification of Th phenotype among bulk cTFH (n=42), HKU1 (n=36), OC-43 (n=34), 229E (n=27), NL63 (n=21) or SARS-CoV-2 (n=20)-specific Tmem. (E) Representative CCR6/CXCR3 expression on bulk (black) or HKU1 S-specific (red) Tmem and cTFH. (F) Paired comparison of the frequency of CCR6^+^CXCR3^-^ cells among hCoV or CoV-2 S-specific Tmem and cTFH populations among responding subjects. HKU, n=34; OC43, n=34; 229E, n=21; NL63, n=18; CoV-2, n=13. Statistics assessed by Wilcoxon test. ***p<0.001, **p<0.01, *p<0.05

Interestingly, hCoV responses comprised a greater proportion of the cTFH population compared to the Tmem compartment in a paired analysis (p<0.002 for all hCoV antigens), with some donors exhibiting a greater than 9-fold enrichment of hCoV-specific cells in the cTFH gate (data for HKU1 shown in Figure 3B). Similar to Tmem, antigen-specific cTFH were highly enriched for a CCR6^+^CXCR3^-^ phenotype (Figure 3C-D). The phenotypes of HKU1- and CoV-2-specific cTFH in SARS-CoV-2-uninfected donors are consistent with phenotypes previously described in COVID-19 convalescent subjects^29^. Comparison of antigen-specific cTFH and Tmem cells revealed a significant enrichment of the CCR6^+^CXCR3^-^ phenotype among cTFH, including the CoV-2 cross-reactive population (Figure 3E-F). These data suggest that while hCoV memory is broadly observed among both Tmem and cTFH subsets, the frequency and phenotype of these responses are, to a degree, subset-specific.

### CoV-2 cross-reactive T cells correlate with HKU1 T cell memory

It is currently unclear whether CoV-2 cross-reactive T cell responses arise primarily from hCoV memory or reflect cross-reactivity from a broad array of antigen specificities^18,25^. Among the cohort, subjects with CoV-2 cross-reactive CD4 T cell responses frequently exhibited memory responses to multiple hCoVs (Figure 4A). We assessed the relationship between the frequency of CoV-2 and hCoV memory responses and found significant correlations only between βCoV and CoV-2 cross-reactivity (p=0.006 for HKU1, p=0.018 for OC43; Figure 4B). This association is consistent with a greater sequence homology among βCoV strains (CoV-2, HKU1 and OC43) compared to the αCoV 229E and NL63^42^.^42^ Among the subset of donors with cross-reactive responses, only HKU1 memory correlated with CoV-2 cross-reactivity (p=0.030, Figure 4B). Interestingly, while almost all individuals with CoV-2 cross-reactive responses exhibited HKU1 and OC43 memory, the converse was not observed. Indeed, individuals with relatively similar patterns of hCoV reactivity could exhibit notably different CoV-2 reactivity (Figure 4C). There was no significant association of demographic characteristics among individuals with or without CoV-2 cross-reactive responses, although the cross-reactive group did exhibit a greater representation of women compared to those without cross-reactivity (p=0.06, Supplemental Figure 3C-D). Given the association between HKU1 and CoV-2 T cell frequencies, we assessed whether a particular phenotype of HKU-specific Tmem was related to the presence or absence of cross-reactive responses, but found no such distinctions (Supplemental Figure 3E).

**Figure 4.**
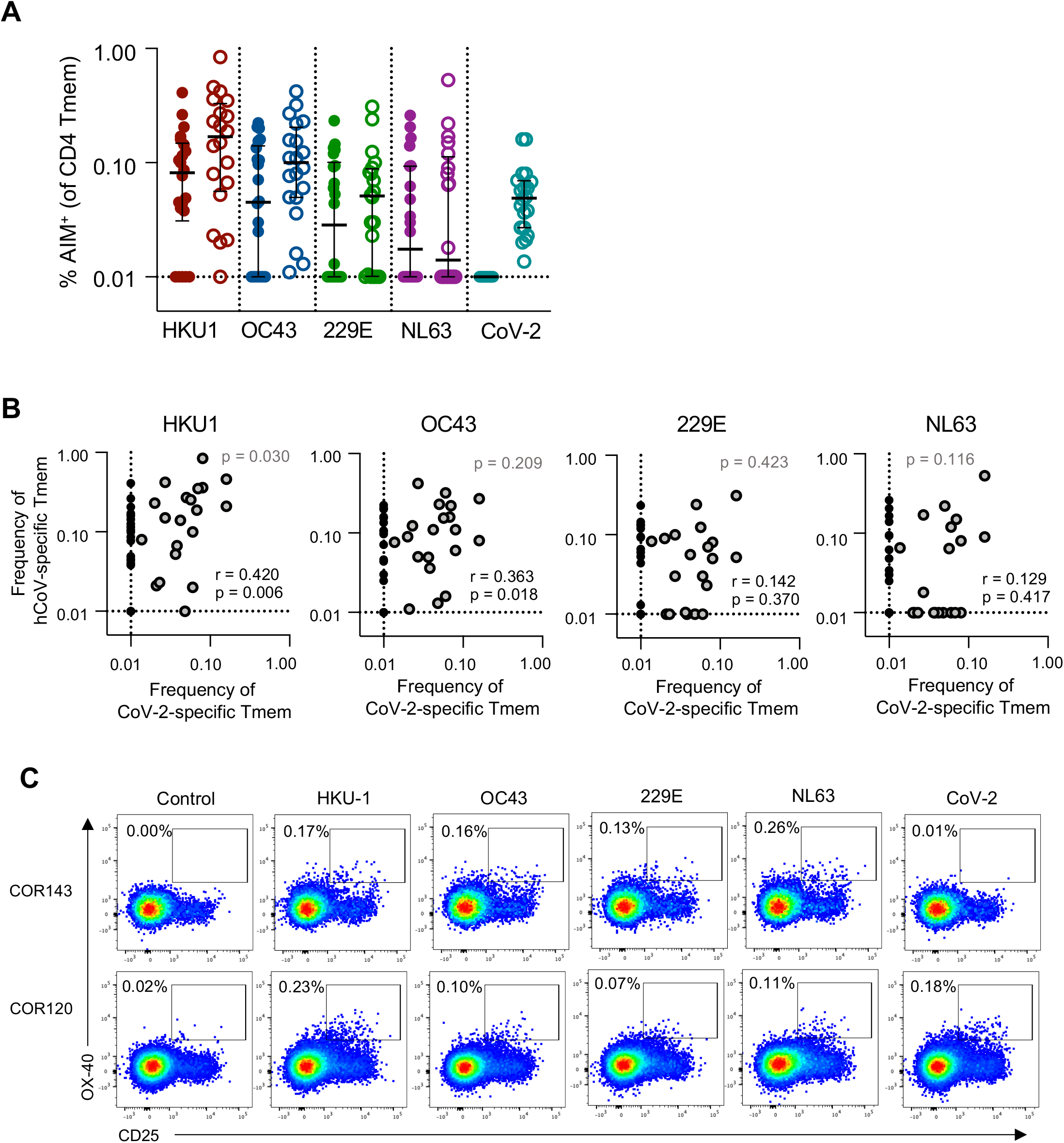
Correlation between HKU1 and CoV-2 cross-reactive Tmem responses. (A) Frequency of S-specific Tmem for each antigen (n=42). Lines indicate median. Values represent background subtracted responses; frequencies below 0.01% after background subtraction were assigned a value of 0.01%. Data points are segregated and coded as individuals without (closed circles, n=22) or with (open circles, n=20) CoV-2 cross-reactive responses. (B) Spearman correlation between the frequency of CoV-2 and hCoV S-specific Tmem (n=42, black text). Correlation p value among individuals with CoV-2 responses >0.01% (n=20, grey dots) is shown in grey text. (C) Representative staining of two healthy donors with S-specific responses to all four hCoV antigens but differential responses to CoV-2 S.

### Cellular and humoral immune memory to hCoV are maintained independently

Studies of convalescent COVID-19 cohorts have demonstrated a strong correlation between SARS-CoV-2-specific cTFH, memory B cells, binding IgG and serum neutralization^19,29,32^, as expected from a coordinated acute immune response. To assess whether such associations are maintained in long-term hCoV immunity, we explored correlations between antibody and T cell responses across the cohort. Surprisingly, there was no relationship for any antigen between plasma IgG endpoint titre and the frequency of either S-specific CD4 Tmem or cTFH (p>0.05 for all; data for HKU1 and NL63 shown in Figure 5A). To gain greater insight into the coordination of cellular and humoral hCoV memory, we undertook an in-depth interrogation of immunity to NL63, which shares use of the cellular entry receptor ACE2 with SARS-CoV and CoV-2, and therefore likely has similar tissue tropisms in vivo.

**Figure 5.**
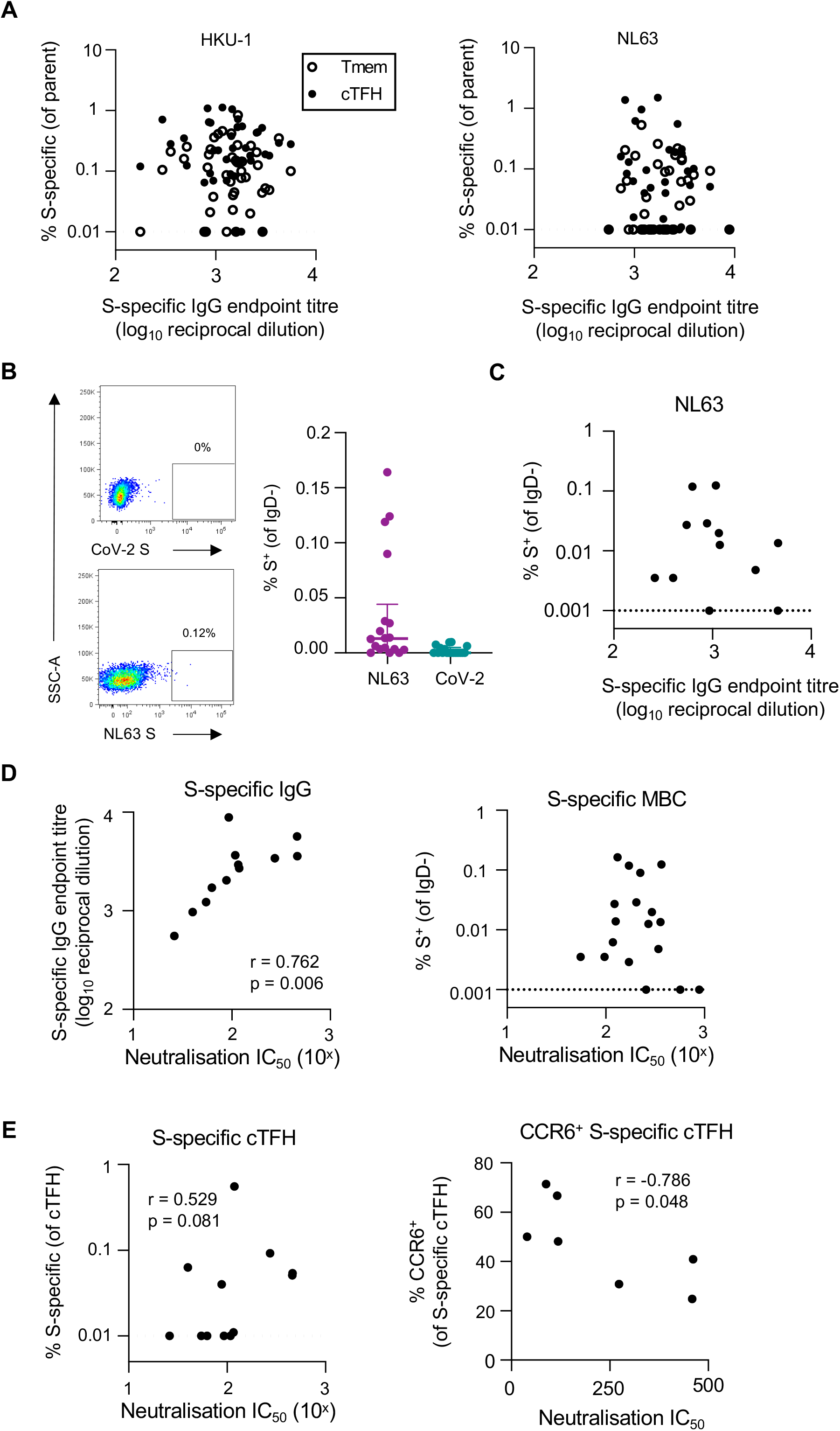
Relationship between serologic, T cell and B cell hCoV memory. (A) Spearman correlation between HKU1 or NL63 S-specific IgG and the frequency of antigen-specific Tmem or cTFH (n=42). Representative staining of IgD-B cells with NL63 or CoV-2 probes and quanification of NL63 and CoV-2 S-specific MBC (n=18). (C) Spearman correlation of NL63 S-specific MBC and plasma binding IgG titres (n=18). MBC frequency was assigned a minimum value of 0.001%. (D) Spearman correlation between plasma NL63 neutralization activity and NL63 S-specific IgG titres or MBC (n=12). (E) Spearman correlation between NL63 neutralization activity and either total NL63 S-specific cTFH or the frequency of CCR6+ antigen-specific cTFH.

To assess S-specific MBC and quantify plasma neutralising activity, NL63 S-specific memory B cell (MBC) probes were generated as described previously^29^, and a novel NL63 pseudovirus-based neutralization assay was performed with 293T cells stably expressing hACE2 as targets. MBC specific for NL63 and CoV-2 S were detected infrequently among the cohort, particularly in comparison to the frequency of CoV-2 S-specific MBC previously reported among COVID-19 convalescent donors^29^ (Figure 5B). Accordingly, the frequency of NL63 S-specific MBC did not correlate with plasma NL63 binding IgG titres (Figure 5C). Plasma neutralising activity against NL63 pseudovirus was detected among all donors tested, with a median IC_50_ of 100.7 (n=12, IQR 56.6-234.6). Neutralising activity strongly correlated with NL63 S-specific antibody titres (p=0.006) but was not associated with NL63 S-specific MBC frequencies (Figure 5D). We did, however, observe a trend toward a positive correlation of neutralization with NL63 S-specific cTFH responses (p= 0.081; Figure 5E). Given our prior observation that CCR6^+^ CoV-2 S-specific cTFH responses were inversely associated with neutralizing antibodies after COVID-19^29^, we assessed the correlation between NL63 neutralising activity and NL63 S-specific cTFH phenotype (for donors with NL63 S-specific cTFH responses, n=7). Interestingly, the frequency of CCR6^+^ cTFH again negatively correlated with plasma neutralization activity (p=0.048; Figure 5E), although the small sample size is a caveat of this analysis.

### Enrichment of HKU1 and NL63 S-specific T cells in lung-draining lymph nodes

Although assessment of hCoV immunity has been primarily limited to peripheral blood, studies suggest that repeated infections with respiratory viruses can seed long-lived memory T cell responses in lung and lung-draining lymph nodes (LDLN)^28,43^.^28,43^ We therefore assessed the frequency of HKU1, NL63 and CoV-2 S-specific CD4 T cell responses in matched LDLN (n=5) and lung samples (n=6) from a human tissue biobank (gating in Supplementary Figure 5). We detected robust HKU1 and NL63 responses within the CD45RA^-^ CD4 T cell population of LDLN (Figure 6A). Given the higher levels of background T cell activation in SLO compared to peripheral blood, we validated the specificity of the hCoV responses by confirming that antigen stimulation also drove expression of CD154 on OX-40^+^ cells (Figure 6A). Among the 5 donors studied, the median frequency of HKU1 and NL63 S-specific Tmem was 1.2% (range 0.12-2.19) and 1.12% (range 0.31-4.04), respectively (Figure 6B). Reactivity to CoV-2 S was substantially lower, with a median of 0.07% (range 0.01-0.99). Similar antigen-specific responses were observed within the CD4^+^CD45RA^-^CXCR5^+^ population (Figure 6B). There was limited to no evidence of ongoing hCoV S-specific GC TFH activity among the samples (data not shown). In contrast to the high frequencies of hCoV-specific CD4 T cells in LDLN, we found only modest hCoV reactivity among lung-derived CD4 T cells (Supplementary Figure 6A-B). These data are consistent with reports that tissue resident T cells (T_RM_) in the lung are relatively short-lived compared to other tissues^44^. Furthermore, the majority of AIM^+^ cells did not exhibit a CD69^+^CD103^+^ phenotype, suggesting they are unlikely to represent bona fide lung-resident T cells^45^ (Supplementary Figure 6C). Similarly, we found little evidence for the presence of NL63 or CoV-2 cross-reactive MBC in either the LDLN or lung tissues (Supplementary Figure 6D-E).

**Figure 6.**
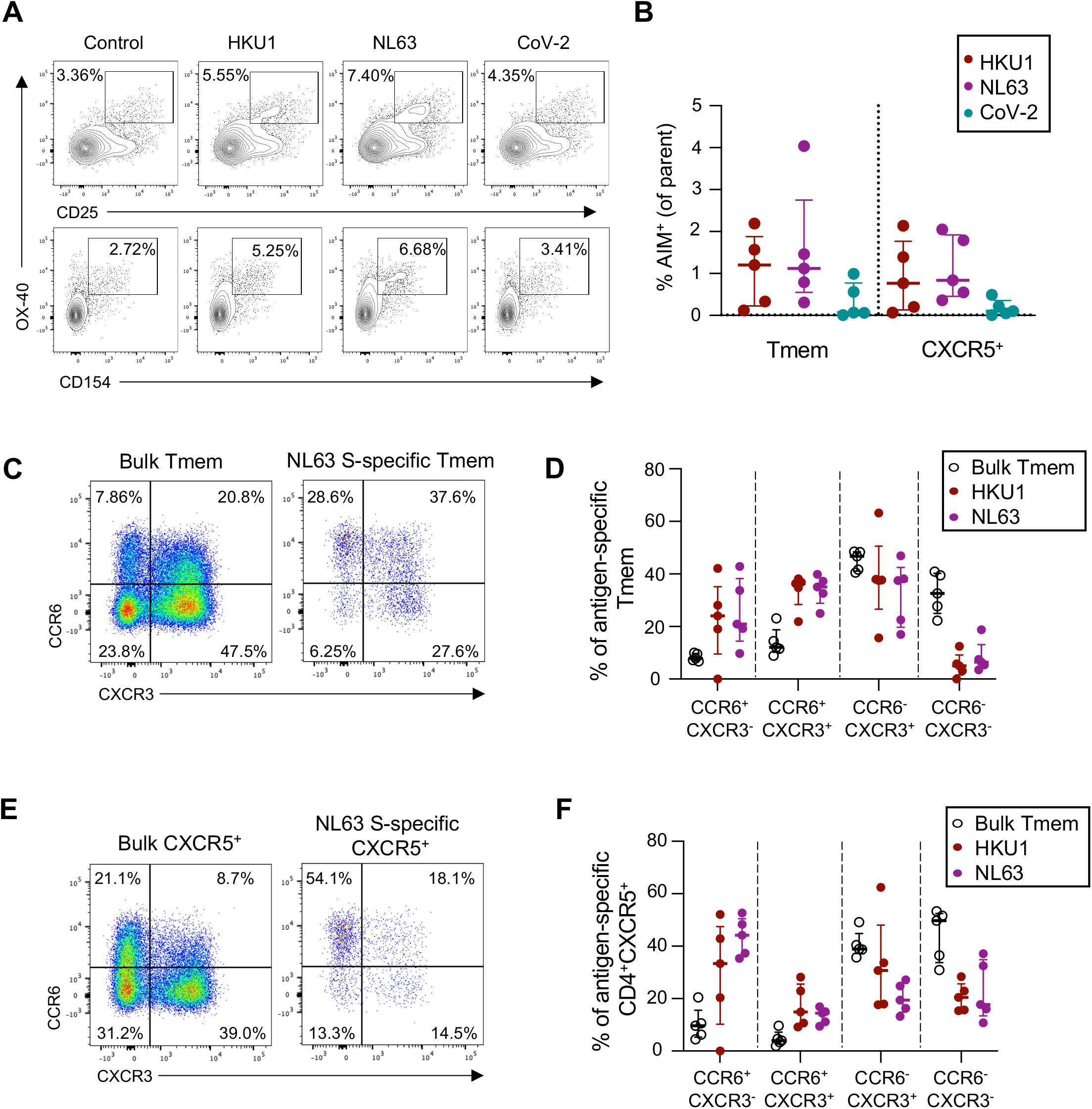
CD4 T cell hCoV memory in human lung draining lymph nodes. (A) Representative staining of AIM and CD154 expression following stimulation with HKU1, NL63 or CoV-2 S among lung-draining lymph node cell suspensions. (B) Frequency of hCoV or cross-reactive CoV-2 responses among Tmem or CD4^+^CXCR5^+^ populations (n=5). (C-E) Representative staining (C, E) and quantification (D, F) of CCR6 and CXCR3 expression on Tmem (C, D) or CXCR5^+^ (E, F) S-specific cells (n=5).

It has been speculated that the dominant CCR6^+^ phenotype of CoV-2-specific CD4 T cells may reflect preferential homing of these cells to the lung^32^.^32^ We therefore compared the CCR6/CXCR3 phenotypes of hCoV-specific T cells in LDLN to the peripheral blood obtained from the unmatched healthy adult cohort presented in Figures 2-3. After adjustment for baseline activation, we found that LDLN-derived hCoV S-specific Tmem exhibited a predominately CXCR3^+^ phenotype, with a substantial population of CCR6^-^CXCR3^+^ cells (median 37.7% for HKU1, 37.5% for NL63; Figure 6C-D). In contrast, only 10.8% and 11.2% of circulating HKU1 and NL63 S-specific Tmem among the blood donor cohort were CCR6^-^CXCR3^+^ (Figure 2D). As we previously observed for cTFH in the periphery, CXCR5^+^ hCoV-specific T cells in the LDLN remained more likely to express CCR6 than their Tmem counterparts (Figure 6E-F). Nevertheless, LDLN-derived hCoV-specific CXCR5^+^ cells were enriched for CXCR3 expression (median 30.7% CCR6^-^CXCR3^+^ for HKU1, 19.4% for NL63) compared to the phenotypes observed among peripheral cTFH (median 6.6% CCR6^-^CXCR3^+^ for HKU1, 9.8% for NL63; Figure 6E-F, Figure 3D). Collectively, these data suggest either differential retention or formation of CXCR3^+^ hCoV S-specific CD4 T cells in LDLN compared to peripheral blood.

## Discussion

Despite periodic re-infection, most adults experience only mild or asymptomatic hCoV infection, suggesting the presence of at least partially protective immune memory. We find that, in addition to near-universal plasma antibody reactivity to hCoV, memory T cell responses to both α- and βCoV are widespread. In contrast, the relatively modest neutralization activity against NL63 and low frequencies of S-specific MBC suggest that sterilizing humoral immunity is likely absent. Instead, additive contributions of multiple arms of adaptive immunity, in particular anti-viral T cell responses, may underpin protection.

While several studies have quantified hCoV-specific T cell responses in adult cohorts^24,25,27^, it was unclear whether the different viruses would elicit phenotypically distinct Tmem or cTFH responses. Together with studies of convalescent SARS-CoV-2-specific T cell responses^29,32^, our data suggest that the CCR6+ phenotype of circulating hCoV-specific CD4 memory cells may be broadly reflective of coronavirus infection in humans. Indeed, AIM-based assays have consistently identified a high proportion of CCR6^+^CXCR3^-^ cells among SARS-CoV-2 S-specific CD4 T cells^29,32^, in spite of low IL-17 production following antigen stimulation^22,29,32^. Interestingly, longitudinal follow-up of COVID-19 convalescent subjects indicated a time-dependent increase in the proportion of CCR6+ S-specific cTFH^33^, suggesting a convergence of phenotypes between CoV-2-specific and hCoV-specific cTFH memory over time. Chemokine receptor expression on CD4 T cells is often used as a surrogate of cytokine expression and Th1/Th2/Th17 function, but these receptors also regulate lymphocyte trafficking to SLO and tissues. While there was little evidence for hCoV S-specific T cell memory in the lung, both HKU1 and NL63 responses were robustly detected in LDLN. The enrichment of CXCR3^+^ hCoV T cell responses in LDLN compared to peripheral blood suggests a potential involvement of CXCR3 expression in recruitment or retention of these cells out of the circulation. These data are consistent with observations in other respiratory infections, where CXCR3 mediates lung trafficking of antigen-specific CD4 T cells^46,47^. Future studies will be required to address the role, if any, for these cells in contributing to protection from re-exposure to CoV infection.

Consistent with other cohorts^17,19^, we find evidence for CoV-2 cross-reactive CD4 T cells in uninfected donors. In vitro expansion of CoV-2 cross-reactive T cell clones has demonstrated the potential for shared specificity with all hCoV^18,24^. However at a cohort-wide level, we find the frequency of CoV-2 cross-reactive cells correlates most strongly with HKU1 memory, although no immediate immunological or demographic features distinguish HKU1-reactive individuals with or without cross-reactive CoV-2 responses. Larger population-based studies will be required to determine any associations between particular HLA class II alleles and cross-reactive CD4 responses. Although it has been speculated that pre-existing cross-reactive T cell immunity could be beneficial in the context of SARS-CoV-2 vaccines^23^, it should be noted that only CXCR3^+^, but not CCR6^+^, cTFH responses appear to correlate with neutralizing antibody titres during COVID-19 convalescence^29,31,48^. While recall of the CCR6^+^ cTFH could induce expression of CXCR3, currently available evidence suggests the highly CCR6-biased responses to hCoV may not be beneficial in the context of vaccination or re-exposure.

Overall, these data clarify the characteristics of long-term immunity to endemic coronaviruses, which have comparable magnitudes and share phenotypic features of S-specific antibody and T cell memory across all four hCoV. Insight into the homeostatic maintenance of hCoV immunity is likely to provide a preview of long-term CoV-2-specific immunity established in the population after vaccination or wide-spread infection.

## Methods

### Subject recruitment and sample collection

SARS-CoV-2 uninfected controls were recruited as part of a previous COVID-19 study^29^, and relevant demographic characteristics are indicated in Figure 1A. For all participants, whole blood was collected with sodium heparin anticoagulant. Plasma was collected and stored at −80 ° C, and PBMCs were isolated via Ficoll Paque separation, cryopreserved in 10% DMSO/FCS and stored in liquid nitrogen. The study protocols and sample use were approved by the University of Melbourne Human Research Ethics Committee (#2056689) and all associated procedures were carried out in accordance with the approved guidelines. All participants provided written informed consent in accordance with the Declaration of Helsinki.

The use of tissue samples from human donors was approved by The University of Melbourne Human Research Ethics Committee (#1954691) and all associated procedures were carried out in accordance with approved guidelines. Tissues were collected from 6 donors: male, age 40-50, brain death; female, 30-40, brain death; male, 30-40, circulatory death; male, 50-60, brain death; female, 60-70, brain death; female, 50-60, brain death. Tissues were passed through 70µM filters and homogenised into single cell suspensions, which were subsequently cryopreserved in 10% DMSO/FCS.

### Expression of coronavirus antigens

A set of trimeric, pre-fusion stabilised coronavirus S proteins (HKU1, 229E, NL63, OC43, SARS-CoV-2) were generated for serological and flow cytometric assays using techniques previously described^29^. Genes encoding the ectodomain of SARS-CoV-2 S (NC_045512; AA1-1209) with 6 proline stabilisation mutations and furin site removal (Hexapro^49^), the HKU1 S (NC_006577; AA1-1291) and NL63 S (DQ445911.1; AA1-1291) with 2 proline stabilisation mutations (S-2P), were cloned into mammalian expression vectors. Plasmids encoding S-2P versions of the ectodomains of OC43 and 229E were kindly provided by Dr Barney Graham, NIH. S proteins were expressed in Expi293 or ExpiCHO cells (Thermofisher) using manufacturer’s instructions and purified using Ni-NTA and size exclusion chromatography. Protein integrity was confirmed using SDS-PAGE.

### ELISA

Antibody binding to recombinant S proteins was determined by ELISA as previously described^29^. Briefly, 96-well Maxisorp plates (Thermo Fisher) were coated overnight at 4°C with 2µg/mL recombinant S, blocked with 1% FCS in PBS, and incubated with plasma dilutions for two hours at room temperature. Plates were washed, incubated with 1:20,000 dilution of HRP-anti-human IgG (Sigma) and developed using TMB substrate (Sigma). Endpoint titres were calculated as the reciprocal serum dilution giving signal 2× background using a fitted curve (4 parameter log regression).

### Flow cytometric detection of hCoV reactive B cells

Probes for delineating NL63 or SARS-CoV-2 S-specific B cells within cryopreserved human PBMC were generated by sequential addition of streptavidin-PE (ThermoFisher) or streptavidin-BV421 (BD), respectively, to trimeric S protein biotinylated using recombinant Bir-A (Avidity). Cells were stained with Aqua viability dye (ThermoFisher). PBMC, lung and lymph node cells were surface stained with the following monoclonal antibodies: CD14-BV510 (M5E2), CD3-BV510 (OKT3), CD8a-BV510 (3GA), CD16-BV510 (3G8), CD10-BV510 (HI10a), SA-BV510 (BD), IgG-BV786 (G18-145), IgD-Cy7PE (IA6-2), and CD19 ECD (J3-119) (Beckman). Cells were washed, fixed with 1% formaldehyde and acquired on a BD LSR Fortessa using BD FACS Diva.

### Flow cytometric detection of antigen-specific CD4 T cells

Cryopreserved human PBMC were thawed and rested for four hours at 37°C. Cells were cultured in 96-well plates at 1-2×10^6^ cells/well and stimulated for 20 hours with 2µg/mL of recombinant S protein from HKU1, NL63, 229E, OC43 or SARS-CoV-2. Selected donors were also stimulated with SEB (1μg/mL) as a positive control. Following stimulation, cells were washed, stained with Live/dead Blue viability dye (ThermoFisher), and a cocktail of monoclonal antibodies. PBMC were surface stained with the following monoclonal antibodies: CD3 BUV395 (SK7), CD45RA PeCy7 (HI100), CD20 BUV805 (2H7), CD154 APC Cy-7 (TRAP-1), CCR7 Alexa Fluor 700 (150503) (BD Biosciences), CD27 BV510 (M-T271), CD4 BV605 (RPA-T4), CD8 BV650 (RPA-T8), CD25 APC (BC96), OX-40 PerCP-Cy5.5 (ACT35), CD69 FITC (FN50), CD137 BV421 (4B4-1), CXCR3 PE Dazzle (G025H7), CCR6 BV786 (G034E3) (Biolegend), and CXCR5 PE (MU5UBEE, ThermoFisher). Monoclonal antibody staining for lung and lymph node cells included: CD45RA PeCy7 (HI100), CD20 BUV805 (2H7), CD154 APC Cy-7 (TRAP-1), EpCam BV711 (EBA-1), CD103 BUV395 (Ber-ACT8) (BD Biosciences), CD3 BV510 (SK7), CD4 BV605 (RPA-T4), CD8 BV650 (RPA-T8), CD25 APC (BC96), OX-40 PerCP-Cy5.5 (ACT35), CD69 FITC (FN50), PD-1 BV421 (EH12.217), CXCR3 PE Dazzle (G025H7), CCR6 BV786 (G034E3) (Biolegend), and CXCR5 PE (MU5UBEE, ThermoFisher) Cells were washed, fixed with 1% formaldehyde and acquired on a BD LSR Fortessa using BD FACS Diva.

### NL63 pseudovirus neutralisation assay

HIV-based lentivirus particles pseudotyped with S from NL63 were generated based on a previously published protocol^50^. Lenti-X 293T cells (TakaraBio) were co-transfected with a lentiviral backbone plasmid expressing Luciferase-IRES-ZsGreen (BEI Resources; NR-52948), helper plasmids encoding HIV Tat, Gag-Pol and Rev (BEI Resources; NR-52948) and a pseudotyping plasmid encoding native NL63 S protein (DQ445911.1). Lenti-X 293T cells were seeded in T175 flasks (18×10^6^ cells/flask) and transfected using lipofectamine (ThermoFisher Scientific) according to manufacturer’s instructions. At 6 hours after transfection, cell culture media was replaced with 36ml of fresh D10 media (DMEM with 10% FCS and 1% PSG). After another 48 hours of incubation, cell culture supernatants containing virions were clarified via centrifugation at 500g for 10 min, filtered through a 0.45µM PES filter and stored at −80 ° C. Infectivity of virions was determined by titration on 293T-ACE2 cells (BEI resources; NR-52511).

For the NL63 pseudovirus neutralisation assay, poly-L-lysine (Sigma Aldrich) coated 96-well white plates (Interpath) were seeded with 293T-ACE2 cells (12,000 cells per well in 60µl). The next day, eight 2-fold serial dilutions of plasma (60µl) were incubated with NL63 pseudovirus (60µl) for 1 hour at 37 ° C (final plasma dilution of 1:20-1:2,560). Plasma-pseudovirus mixtures (100µl) were then added to 293T-ACE2 cells and incubated at 37 ° C for 48 hours. Plasma samples were tested in triplicate, with “virus+cells” and “virus only” controls included to represent 100% and 0% infectivity respectively. After 48 hours, all cell culture media was carefully removed from wells. Cells were lysed with 25µl of passive lysis buffer (Promega), incubated on an orbital shaker for 15 mins and developed with 30µl britelite plus luciferase reagent (Perkin Elmer). Luminescence was read using a FLUOstar Omega microplate reader (BMG Labtech). The relative light units (RLU) measured were used to calculate %neutralisation with the following formula: (‘Virus+cells’ – ‘sample’) ÷ (‘Virus+cells’ – ‘Virus only’) × 100. The half maximal inhibitory concentration for plasma (IC_50_) was determined using four-parameter nonlinear regression in GraphPad Prism with curve fits constrained to have a minimum of 0% and maximum of 100% neutralisation.

### Statistics

Statistical analysis was performed in GraphPad Prism v9. All T cell stimulation data is presented after background subtraction using the unstimulated control. Two group comparisons were performed using the Mann-Whitney test, or the Wilcoxon test for paired data. Correlations were performed using the Spearman test. P values were considered significant if <0.05.

## Data Availability

All data is available from the authors upon reasonable request

## Acknowledgements

The authors would like to thank the study participants and clinical teams for their participation in and assistance with the study. We gratefully acknowledge the generosity of the organ donor families for providing valuable tissue samples. We acknowledge the Melbourne Cytometry Program for provision of flow cytometry services. Funding for this work was provided by the Victorian Government, a Doherty Collaborative Research Award (JJ), the ARC Centre of Excellence in Convergent Bio-Nano Science and Technology (SJK), an NHMRC program grant APP1149990 (SJK), and philanthropic support from the Paul Ramsay Foundation (SJK and AKW). AKW is funded by an NHMRC Investigator grant. JAJ and SJK are funded by NHMRC fellowships.

## Author Contributions

HXT, WSL, AKW, SJK and JAJ designed the study, analysed the results and wrote the manuscript. KMW, HXT, CN, WSL, RE, HGK, TA, AKW and JAJ performed the experiments. RJ, GS, BZW, OY, TT, MLG, HO, RD, AV, LKM, CLG facilitated collection of human tissues. All authors read, edited, and approved the manuscript.

## Supplementary Figures

**Supplementary Figure 1.**
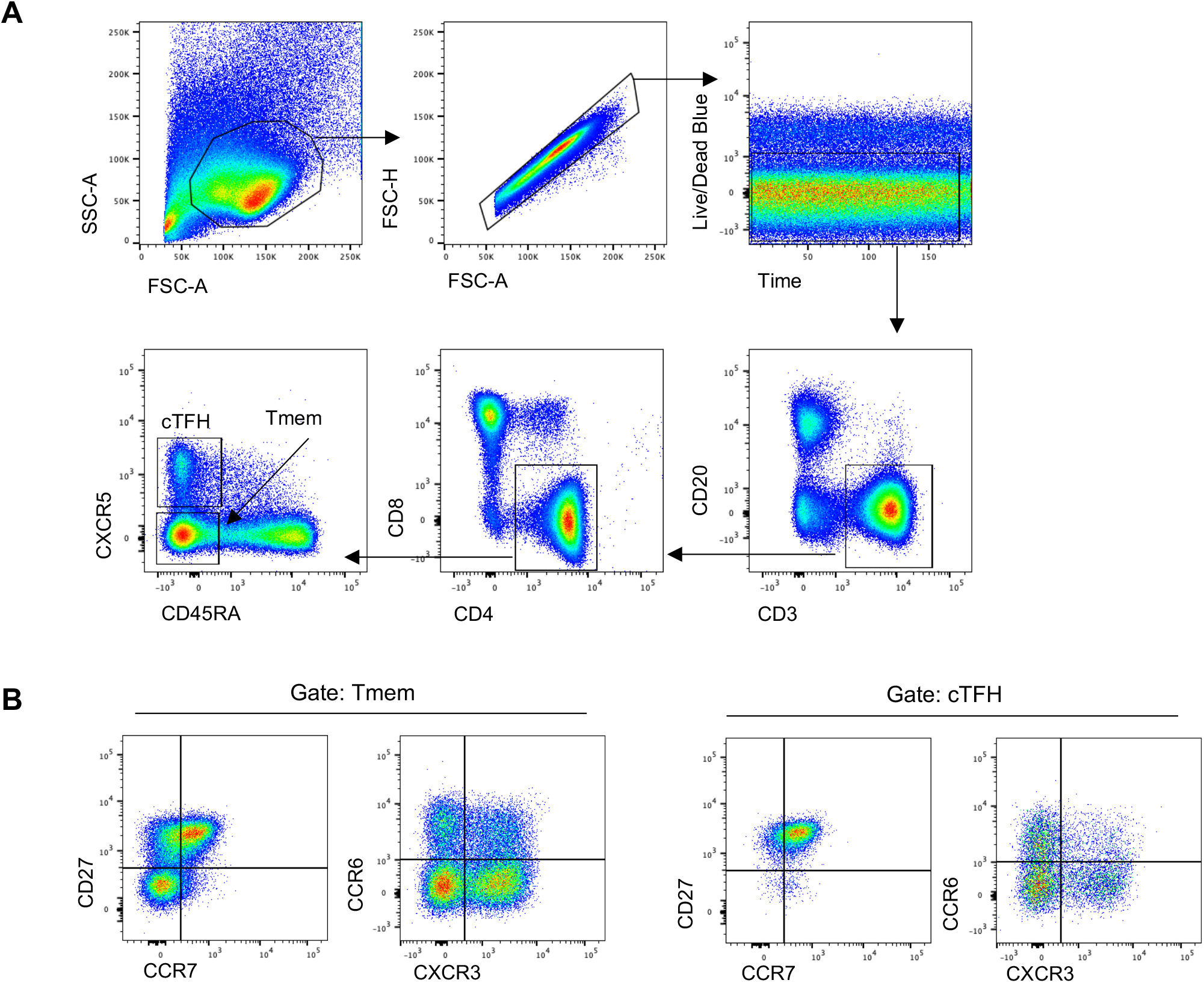
CD4 T cell gating strategy. (A) Lymphocytes were identified by forward and side scatter, followed by doublet exclusion (FSC-A vs FSC-H), and gating on live cells with a consistent fluorescence profile over time. T cells were identified as CD20-CD3+, and CD4+CD8-cells were further defined as cTFH (CXCR5+CD45RA-) or Tmem (CXCR5-CD45RA-). (B) Tmem and cTFH populations were phenotyped using memory markers (CD27 vs CCR7) or chemokine receptors (CCR6 vs CXCR3).

**Supplementary Figure 2.**
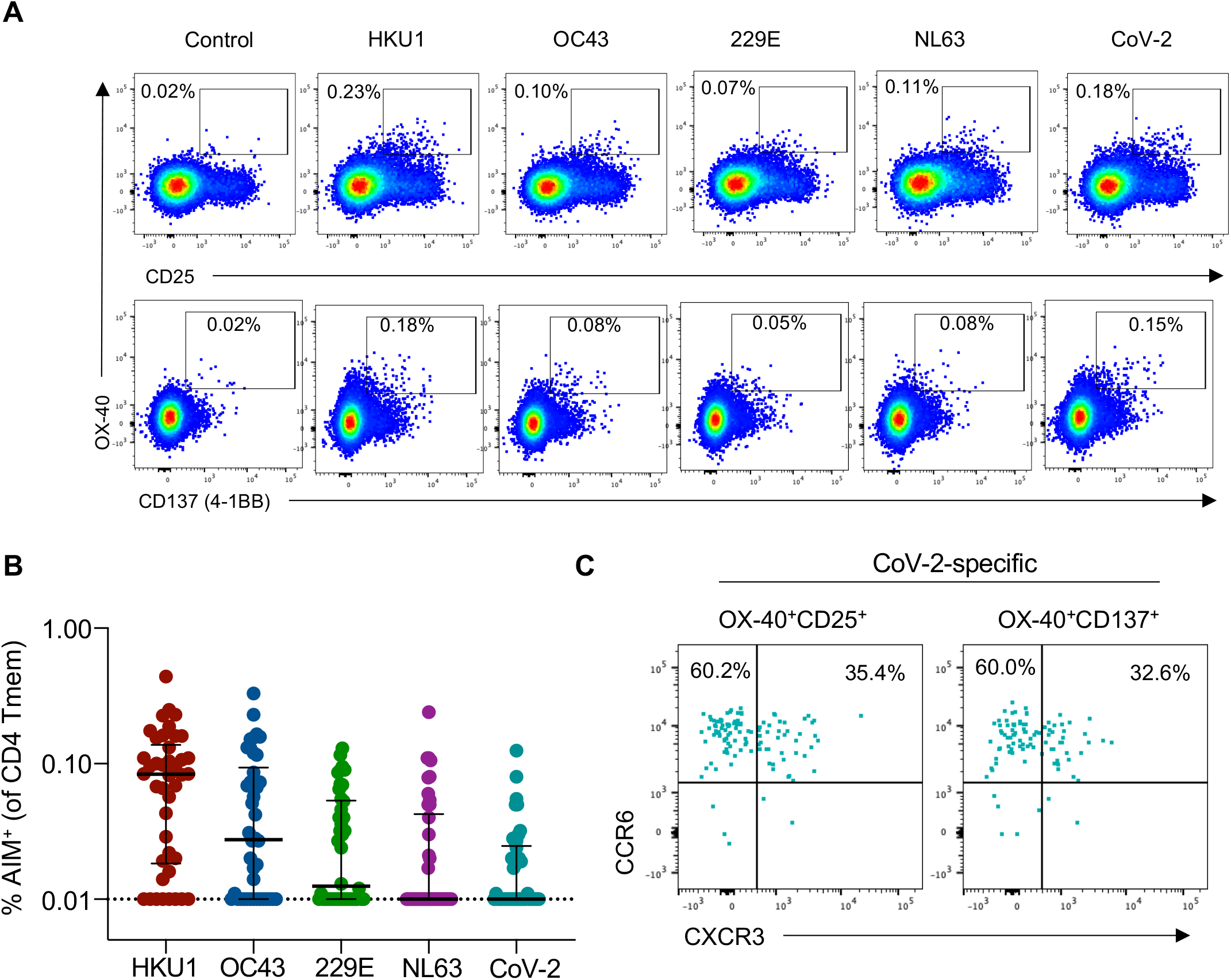
CD4 Tmem responses measured by different AIM combinations. (A) Comparison of AIM marker readouts (OX-40+CD25+ vs OX-40+CD137+) in a single individual for S-specific responses to all four hCoV antigens and SARS-CoV-2 S. (B) Frequency of S-specific Tmem for each antigen (n=42). Bars indicate median. Values represent background subtracted responses; frequencies below 0.01% after background subtraction were assigned a value of 0.01%. Symbols and colours indicate individual donors with SARS-CoV-2 responses >0.01% and are matched to the individuals coded in Figure 2C. (C) Comparison of SARS-CoV-2-specific Tmem CCR6 and CXCR3 phenotype based on identification by OX-40^+^CD25^+^ or OX-40^+^CD137^+^ gates.

**Supplementary Figure 3.**
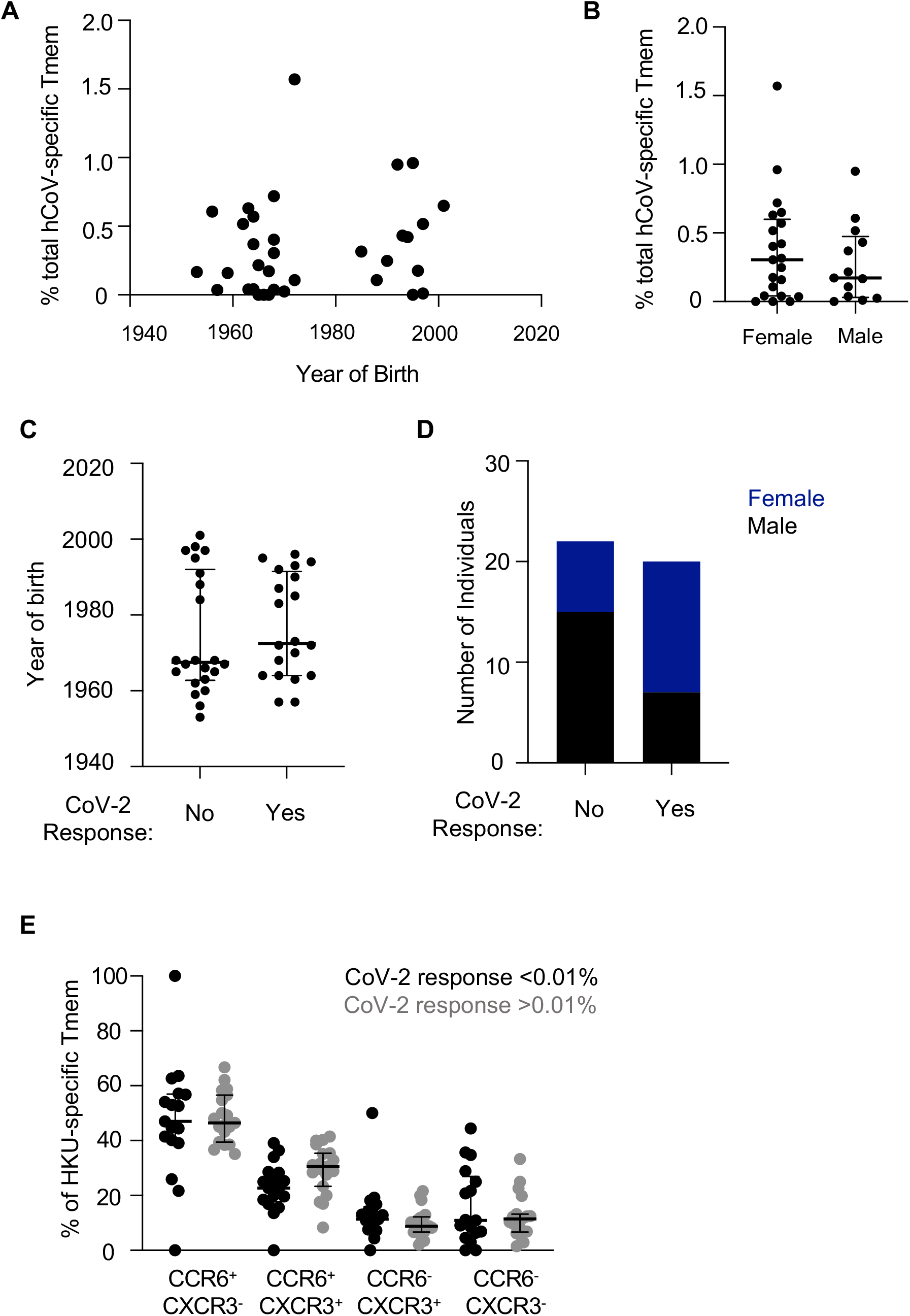
Demographic and immunological characteristics of total hCoV and CoV-2 cross-reactive responses. (A) Spearman correlation of age and total hCoV Tmem frequency (n=42). (B) Total hCoV frequency according to gender (n=42). (C) Age and (D) gender distribution among individuals with no (<0.01%) CoV-2 S-specific CD4 Tmem responses (n=22) or with CoV-2 responses (n=20). (E) Phenotype of HKU1-specific Tmem in individuals without (black, n=17) or with (grey, n=19) cross-reactive CoV-2 responses. Lines and error bars indicate median and interquartile range.

**Supplementary Figure 4.**
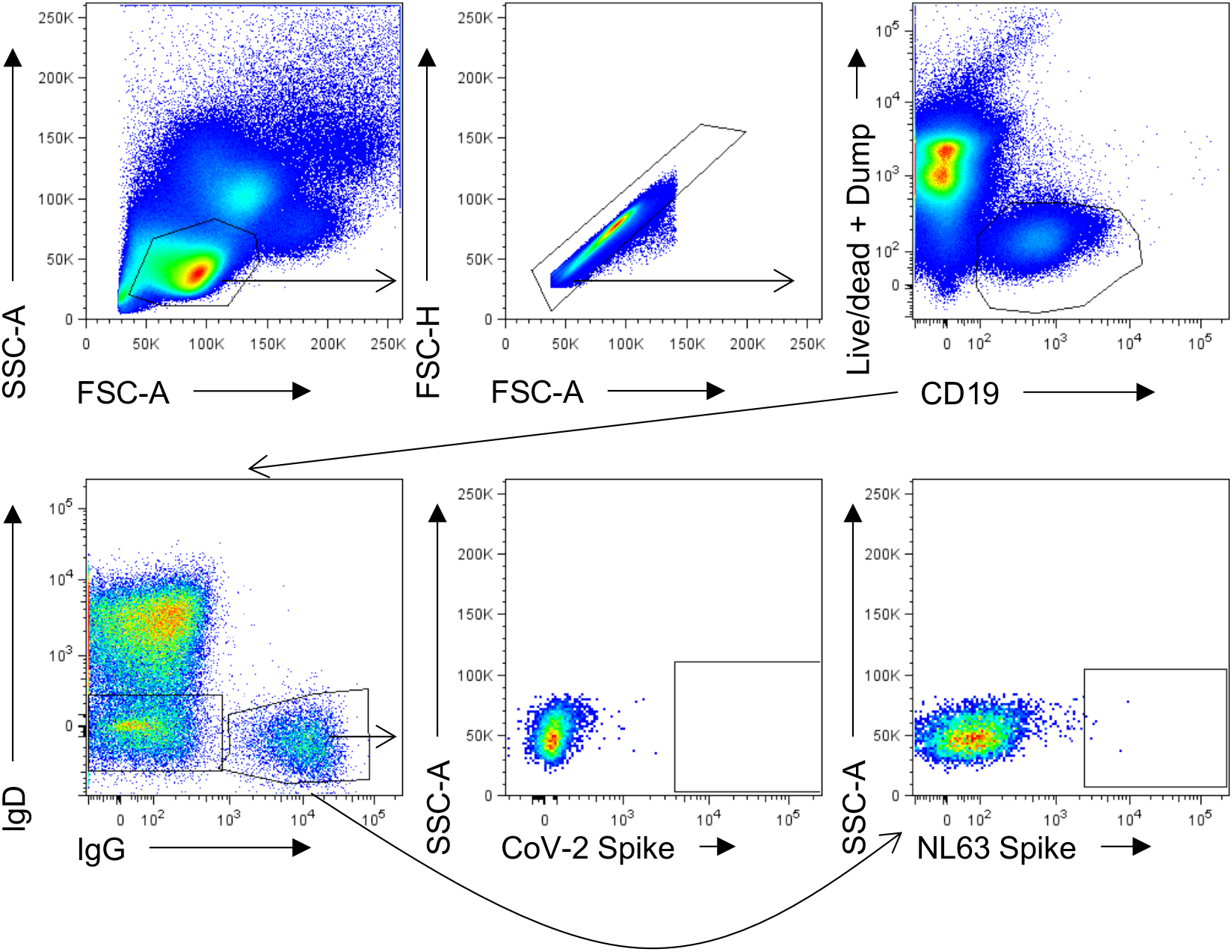
Gating strategy for resolving antigen-specific B cells. Lymphocytes were identified by FSC-A vs SSC-A gating, followed by doublet exclusion (FSC-A vs FSC-H), and gating on live CD19+ B cells. Class-switched B cells were identified as IgD-, IgG+. Binding to SARS-CoV-2 or NL63 spike was assessed.

**Supplementary Figure 5.**
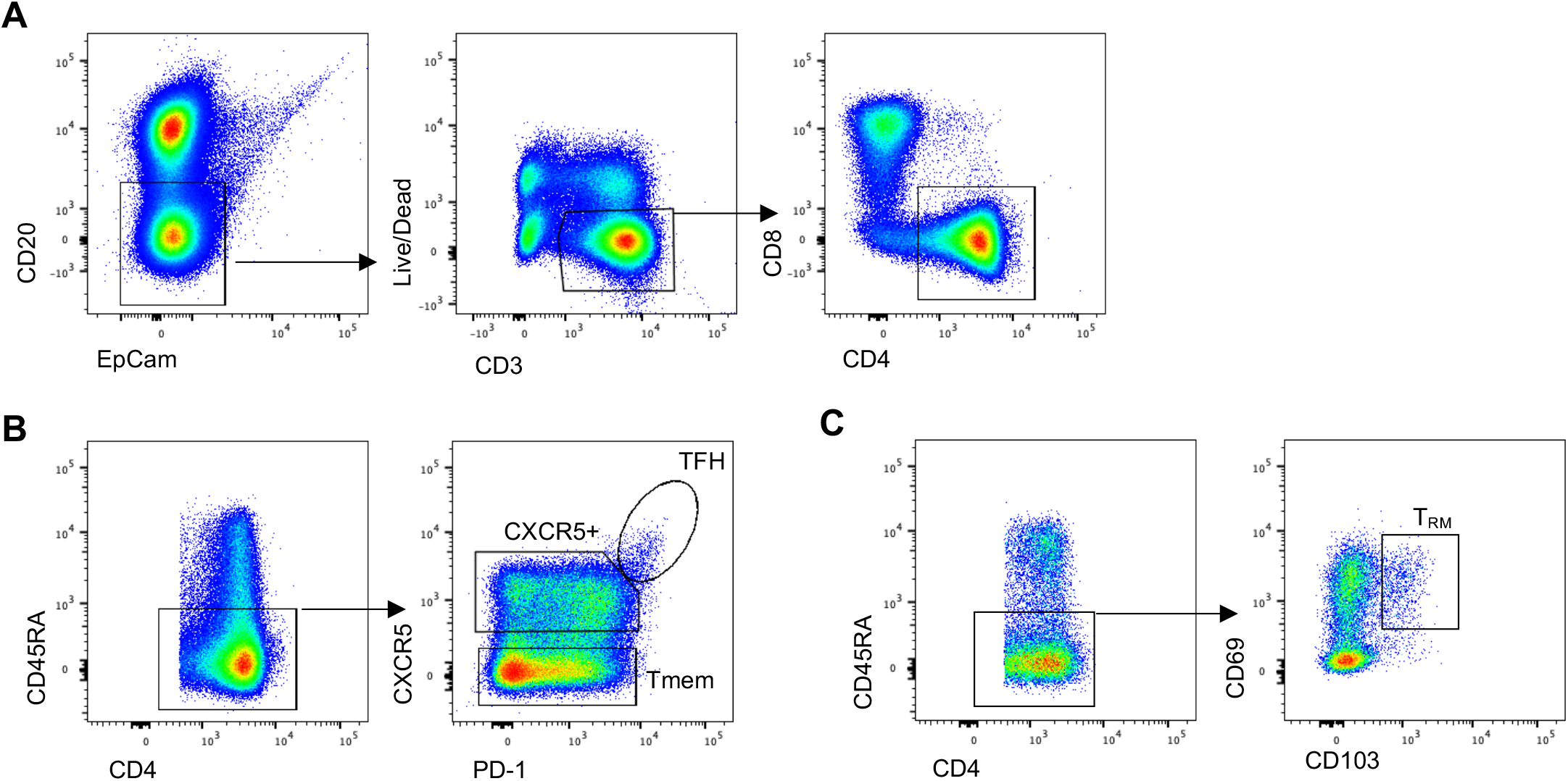
Lymph node and lung CD4 T cell gating strategy. (A) CD4 T cells in lymph node or lung samples were identified as lymphocytes (identified by forward and side scatter, followed by doublet exclusion) with a CD20-EpCam-Live/Dead-CD3+CD8-CD4+ phenotype. (B) Memory CD4 T cell subsets in lung draining lymph nodes were identified as CD45RA-, followed by gating based on PD-1 and CXCR5 expression to identify TFH, pre-TFH and Tmem subsets. (C) In lung samples, memory CD4 T cells were identified as CD45RA-. T_RM_ cells were identified as CD69+CD103+.

**Supplementary Figure 6.**
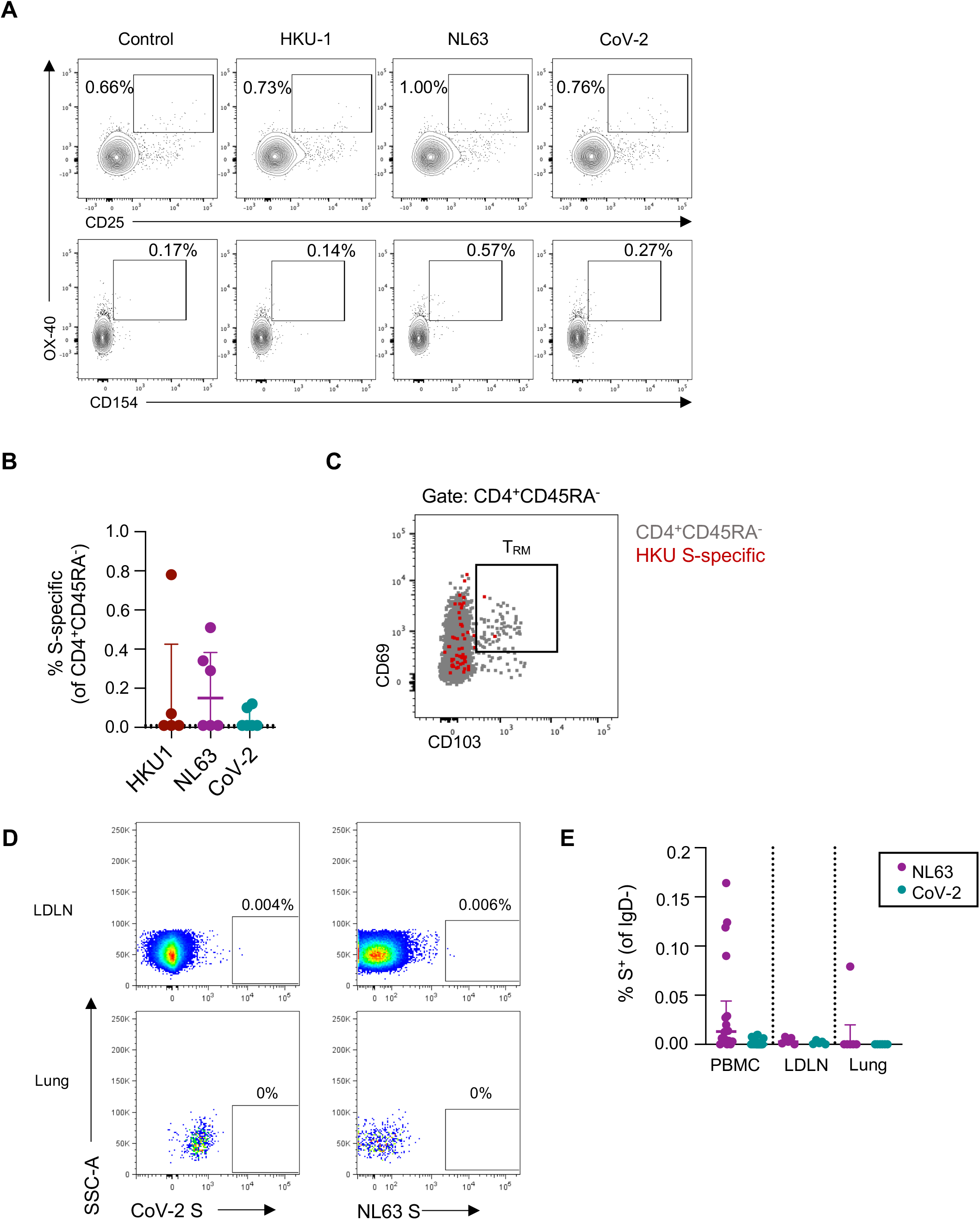
hCoV and CoV-2 T cell and B cell responses in LDLN and lung. (A) Representative responses to HKU1, NL63 or CoV-2 S antigen stimulation among CD4^+^CD45RA^-^ T cells in the lung. (B) Quantification of S-specific T cell responses (n=6 for NL63 and CoV-2, n=5 for HKU1). (C) Expression of CD69 and CD103 on total CD4^+^CD45RA^-^ lung T cells (grey) versus HKU S-specific T cells (red). (D) Representative staining of NL63 and CoV-2 S-specific MBC in LDLN and lung samples. (E) Quantification of S-specific MBC among PBMC samples from the healthy adult cohort (n=18), or human tissue biobank LDLN (n=6) or lung (n=6).

